# Genetic Causes of early onset Ataxia: Experience from the National Sheffield Paediatric Ataxia Centre

**DOI:** 10.1101/2024.07.03.24309227

**Authors:** K Garrard, N Beauchamps, DJA Connolly, S Secker, K E Allen, S Campbell, M Panayi, M Hadjivassiliou, SR Mordekar

**Affiliations:** Biomolecular Sciences Research Centre, Sheffield Hallam University, City Campus, Howard Street, Sheffield, S1 1WB; Sheffield Children’s Hospital, Western Bank, Sheffield, South Yorkshire, S10 2TH; University of Sheffield, Sheffield, South Yorkshire S10 2JF

**Keywords:** Paediatric ataxia, diagnostic genetics, humans, cerebellar ataxia, spinocerebellar ataxia, High-throughput nucleotide sequencing

## Abstract

Early onset cerebellar ataxias are sufficiently distinct in aetiology and disease course from adult onset ataxias to warrant independent evaluation. It has long been assumed that complex (multisystem) ataxias are more frequent in the paediatric ataxia population but the proportion of genetic causes and the makeup of this group of patients has not previously been examined in detail.

Data from 704 patients from the Sheffield Paediatric Ataxia Centre (SPAC) confirms Friedriech’s ataxia as the most common genetic paediatric ataxia (25%) but this is closely followed by *CACNA1A* mutations (18.2%). Pick up rate was higher than for adult populations and recessive and dominant conditions were represented in roughly equal proportions. A large proportion of mutations were only found in a single gene and nearly half of the NGS variants identified (46.7%) were variants of unknown significance (VUS). In total 13.8% of this population had a genetic cause confirmed. This demonstrates the utility of large gene panel testing in the paediatric ataxia population and highlights the need for further research and developments into determination of the pathogenicity of genetic variants.

In conclusion, simple mendelian genetic diseases are responsible for a significant proportion of cases of chronic ataxia in the paediatric cohort.

## Introduction

The word ataxia comes from the Greek “a” without or lacking and “taxis” order. Ataxia describes an abnormal movement which is uncoordinated and inaccurate. It is a marker for cerebellar damage or degeneration and consequently paediatric ataxia represents a collection of heterogeneous disorders with a shared anatomical location for their pathology. Cerebellar damage can manifest as ataxia in different parts of the body leading to; gait instability, dysarthria, axial hypotonia and nystagmus. Cerebellar damage can also lead to executive function difficulties and this is described as cerebellar cognitive affective syndrome. In addition, in the paediatric population, ataxia can present with dyslexia or dyspraxia, making a definite diagnosis of ataxia more difficult than the adult population.

Most large cohort ataxia studies to date have published data exclusively from adult patients or have failed to distinguish between paediatric and adult populations (Hadjivassiliou *et al* 2018, Subramony *et al* 2022). Paediatric ataxia may have a distinct aetiology with a different balance between environmental and innate causes and as such, it is important to examine this group independently. This paper aims to rectify this oversight by examining genetic and diagnostic findings exclusively in a paediatric population. We have assembled data from patients referred to the Sheffield Children’s Hospital for ataxia testing over a ten year period (2009-2019) and used this data to inform future improvements to local diagnostic practices in paediatric ataxia.

Paediatric ataxia only accounts for 6% of all cases of ataxia (Hadjivassiliou *et al* 2018). The prevalence of paediatric and early onset (under 20) ataxia is predicted to be between 14.6-26 per 100,000 (Brandsma *et al* 2019, Mordekar *et al* 2019). Paediatric ataxia can be classified in several different ways; anatomy, aetiology or time of presentation. In this paper we will use time of presentation and cases will be classified as chronic, acute or episodic.

Acute paediatric ataxia is often (more than 50%) caused by intoxication (Mordekar *et al* 2019) and is a common acute presentation to Paediatricians. Other prevalent causes include; post viral cerebellitis, labyrinthitis, acute disseminated encephalomyelitis and Vertebral artery dissection. Intermittent or episodic ataxia (EA) is very rare in children and most cases are due to pathogenic changes in channelopathy genes (Mordekar *et al* 2019).

Most chronic ataxias, childhood and adult onset, are progressive. They can be subdivided by aetiology into developmental, metabolic or genetic.

Ataxia presents differently depending on the age of the patient. Ataxia in the paediatric population, especially that with congenital onset, is more likely to have a genetic than an environmental cause compared to adult onset ataxia. This is principally due to lower risk of exposure to long term environmental toxins (Hadjivassiliou *et al* 2018). As is often the case with genetic disease, there is often multisystem involvement leading to complex phenotypes which can evolve over time. Many of the individual disorders are rare and the phenotypic range may not be well characterised. Other indicators that ataxia may have genetic causes are family history and congenital onset.

A major factor when considering diagnosis of the cause of ataxias in children is balancing the cost of diagnosis against the availability and cost of treatment (Lynch *et al* 2018). Many ataxias, especially genetic ataxias, are rare and as a consequence only have supportive management strategies. With ongoing advances in technology enabling faster and cheaper genetic testing and the potential for repurposing existing drugs (Alfedi *et al* 2019, Perlman 2020) the number of treatable paediatric ataxias is likely to increase, making such decisions easier.

## Materials and methods

This study was registered as a service evaluation project with the Audit department at the Sheffield Children’s Foundation Trust.

The Sheffield Paediatric ataxia centre opened in 2013 based at Sheffield Children’s NHS Foundation Trust. In 2018, it was accredited by Ataxia UK as a paediatric ataxia centre of excellence. Referrals are accepted from secondary care paediatricians from England. Once referred, patients are reviewed by a paediatric neurologist with expertise in ataxia and are taken through the diagnostic pathway for ataxia (see Fig. 1). As part of this, detailed assessment is undertaken to exclude ataxia mimics. Specialist neuroimaging is undertaken and scale for assessment and rating of ataxia (SARA) score is recorded on all children referred and on follow up.

**Fig 1.**
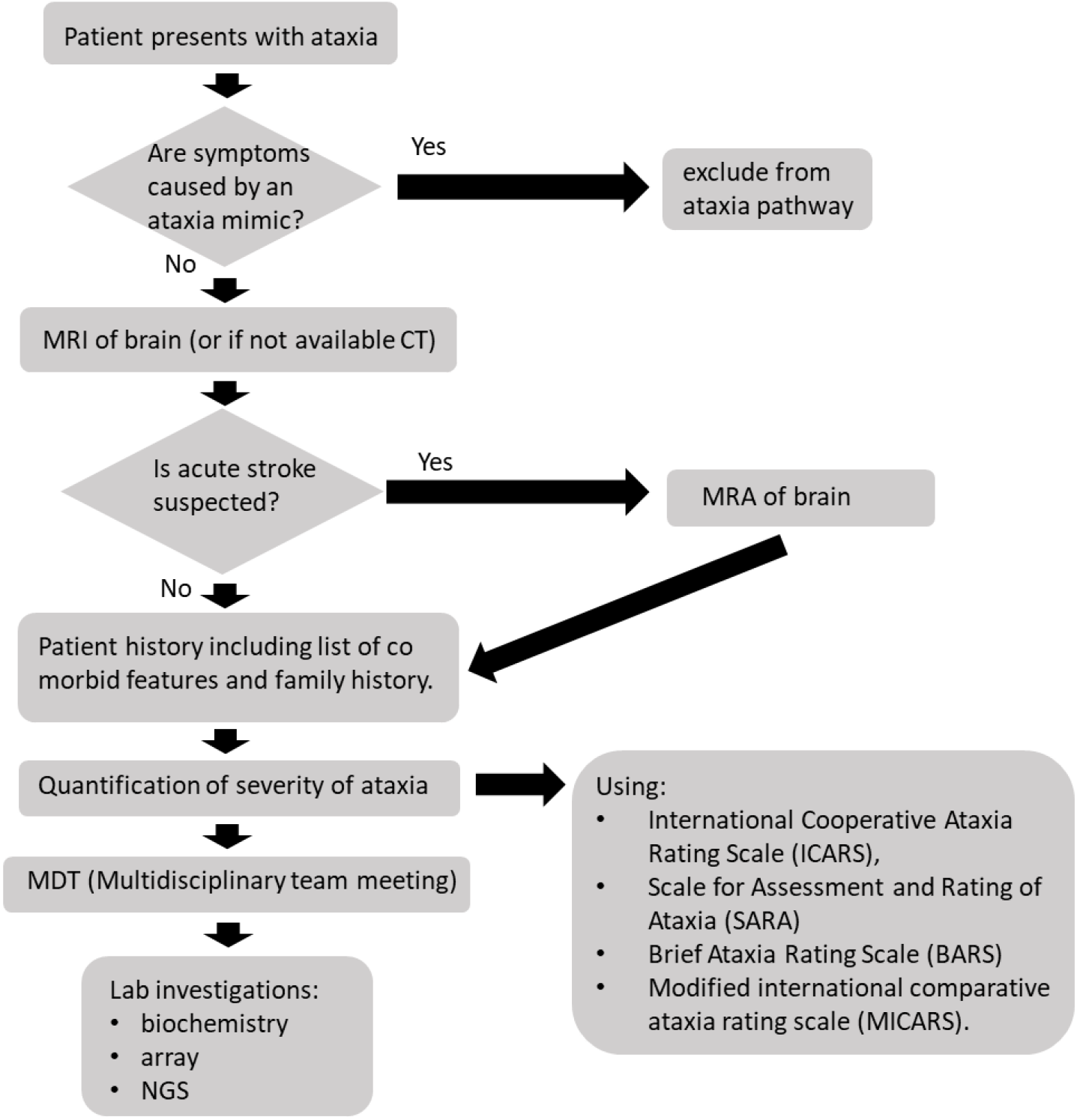
Diagnostic pathway for paediatric ataxia

Following assessment by a multidisciplinary team, Investigations are undertaken and the families are signposted to allied specialty services and local teams to support the child’s management.

### Brain imaging

Magnetic resonance (MR) imaging of the brain has routinely been employed in the evaluation of ataxia in children over the timeframe of this review. All children undergo neuroimaging at referral and then monitoring annually. This is achieved via MRI brain scan with MR spectroscopy of the cerebellum with voxel at the cerebellar vermis and right cerebellar hemisphere using a standard protocol.

If there is an acute presentation then magnetic resonance angiography (MRA) with review of the vertebral and carotid arteries for dissection, diffusion weighted imaging (DWI) to assess for acute stroke and a blood sensitive sequence such as susceptibility weighted imaging (SWI) or gradient echo T2* imaging to look for haemorrhage is included.

Standard imaging for intermittent or chronic ataxias includes axial T2, T1 volume and FLAIR imaging. Comment will be made regarding the supratentorial brain appearances. Assessment for potential atrophy of the brainstem, vermis and cerebellar hemispheres. The radiologist will also assess for focal signal change on T1 and T2 images in the brainstem, cerebellum and dentate nuclei regions. Specific imaging diagnoses are rare in paediatric ataxia (e.g. ARSACS) but may become better recognised as further understanding of the pathologies and genetic disorders is achieved.

### Genetic testing

In the UK, pre-2015 routine genetic testing was only available for a small number of ataxias; principally those associated with triplet repeat expansions; the common spinocerebellar ataxias (SCA1, SCA2, SCA3, SCA6, SCA7), dentatorubral-pallidoluysian atrophy (DRPLA) and Friedreich’s ataxia (FA) or for single gene disorders such as episodic ataxia type 2 (EA2). This approach was not very effective for paediatric ataxia patients as previous studies suggested that most SCAs are not childhood onset and should not be considered in children unless there is a family history (Lynch *et al* 2018). These forms of testing only allowed testing of a single gene at a time, either in parallel, as with the common SCA’s (2,3,4,6 and 7), FA and DRPLA or in series, as with *CACNA1A*, where the gene was screened in two parts, with the most commonly variable exons checked first, followed by the rest of the gene. Over the last decade, improvements in technology and reductions in costs have made it possible for next generation sequencing (NGS) to be utilised in diagnostic settings, allowing many genes to be screened concurrently.

In 2015 an NGS ataxia panel was designed and brought into service by Sheffield Diagnostic Genetics Service (SDGS). It was based on extensive literature searches and OMIM (online mendelian inheritance in man, 1987) searches for single gene disorders with phenotypes which included ataxia. The results were cross referenced against the 100,000 genomes panel app to ensure that genes were clinically appropriate for inclusion. Both adult and childhood onset conditions were included.

Retrospective data for all paediatric ataxia testing between Jan 2010 and Dec 2019 was collected from SDGS. Data was pseudo anonymised and analysed according to the testing carried out. Positive diagnostic rates were determined overall and for each test. Variants identified from NGS analysis were assessed for pathogenicity using the American standards for assessment of pathogenicity (Richards *et al* 2015) and UK guidelines for assessment of pathogenicity (Ellard *et al* 2020). Patients with single pathogenic variants in autosomal recessive genes or variants of uncertain clinical significance in autosomal dominant genes have been classified as “inconclusive” results for the purpose of this report. All testing and reporting was carried out by an accredited diagnostic genetics laboratory in accordance with relevant guidelines and procedures with consent obtained from the parents of the affected children

Diagnostic rates for single gene and NGS testing were compared.

**Table 1:**
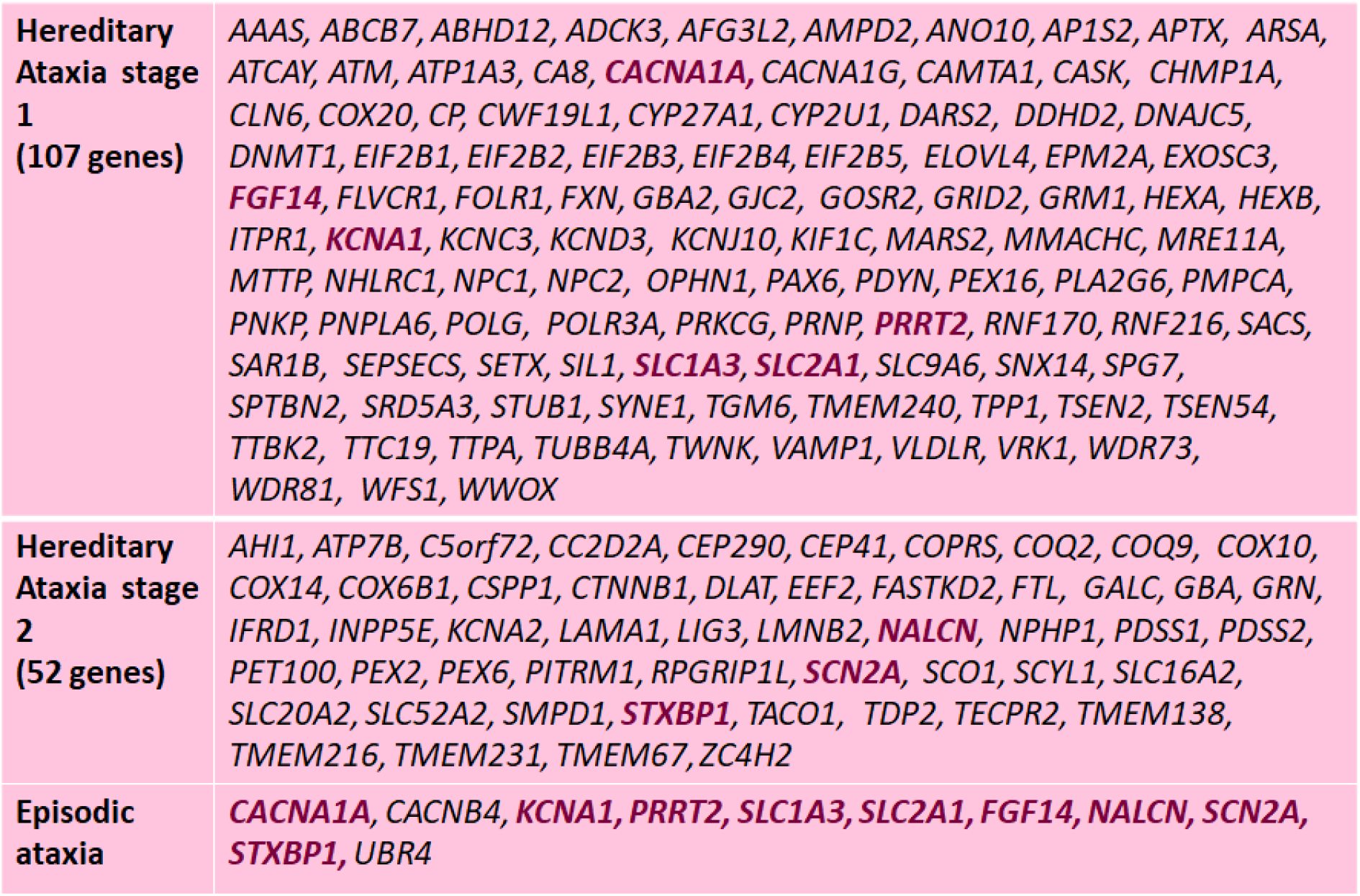
genes contained in the SDGS NGS ataxia panel and its subgroups.

## Results

### Neuro imaging

During the period of this study, 63 patients underwent MRI. 54 of the 63 scanned (86%) had abnormal MRI with structural abnormalities or abnormal MR spectroscopy of cerebellum.

### Genetic testing

704 ataxia patients were referred to SDGS for genetic testing: 219 ataxia single gene,134 ataxia NGS, 166 EA single gene, and 185 EA NGS panel. Genetic diagnoses were identified in 16 (7.3%) single gene ataxia tests and 32 (24%) of ataxia NGS with a further 28 (21%) NGS tests inconclusive. Genetic diagnoses were identified in 32 (19.3%) of single gene EA tests and 17 (9.2%) of EA NGS panels with inconclusive results in a further 8 (4.3%) of NGS tests.

The most common cause of genetic ataxia in our cohort was Friedreich ataxia (11 (25%)), followed by *CACNA1A* pathogenic mutations (8 (18.2%)) and then ataxia telangiectasia (5 (11.4%)) (Fig. 3). The values of the remaining disorders identified are too small to draw conclusions about prevalence within the paediatric ataxic population. Some rare ataxias have specific genetic testing referral pathways and so genetic data may be underrepresented (e.g. ataxia telangiectasia referrals are often sent directly to the West Midlands Regional Genetics Laboratory).

**Fig 2.**
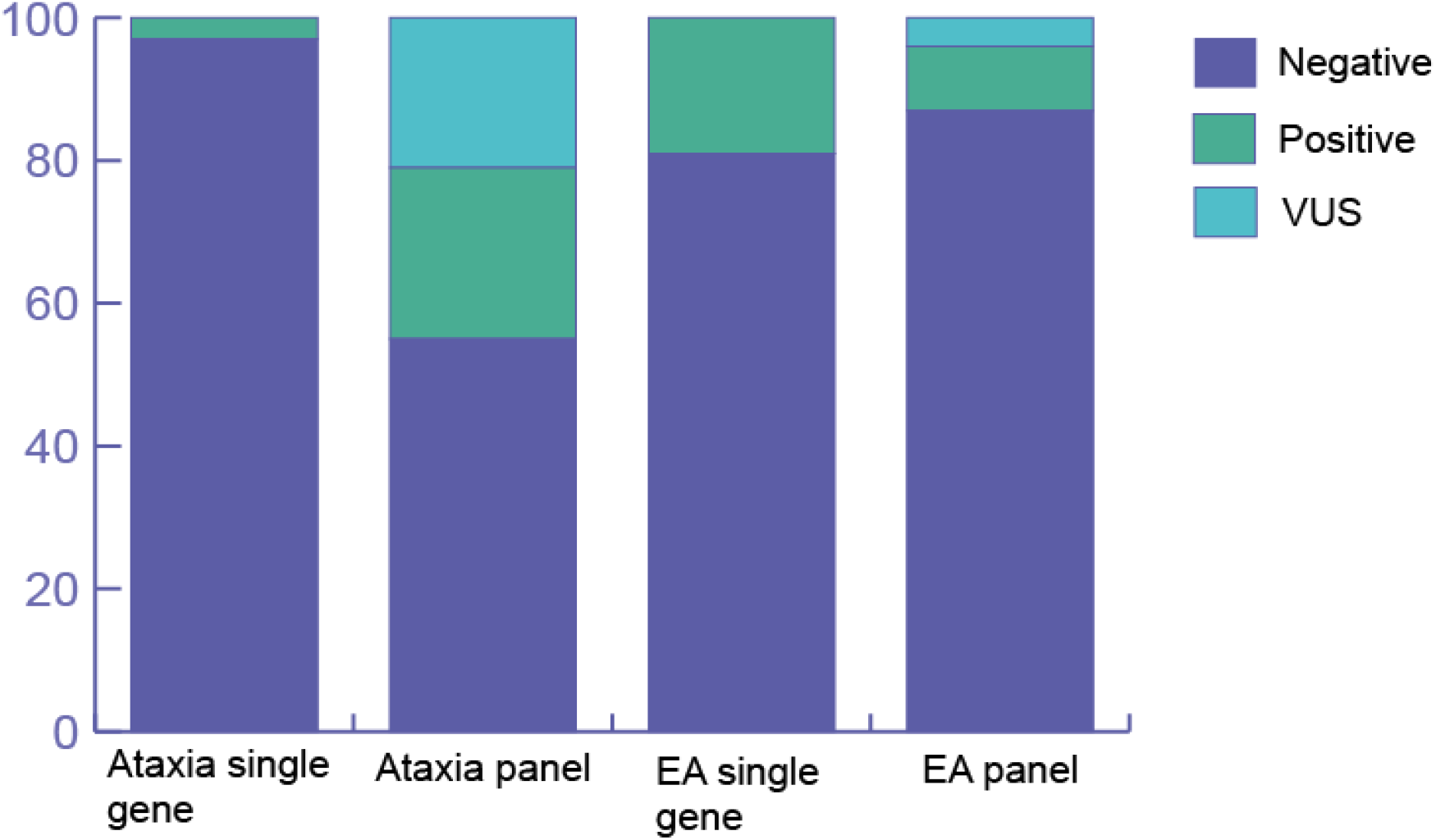
Breakdown of results from ataxia and EA single gene and panel testing as a percentage of totals.

**Fig 3.**
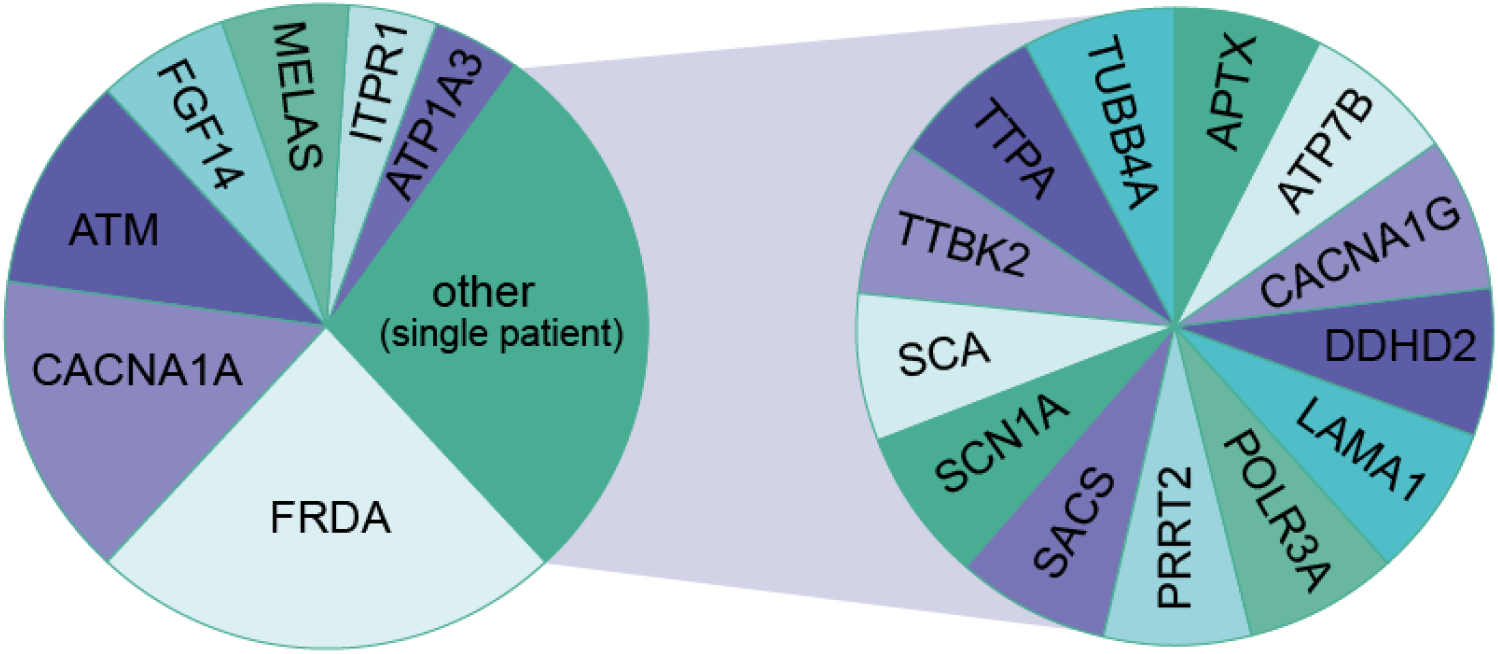
Proportions of genes (single gene and NGS testing) in which pathogenic DNA variants were identified

**Fig 4.**
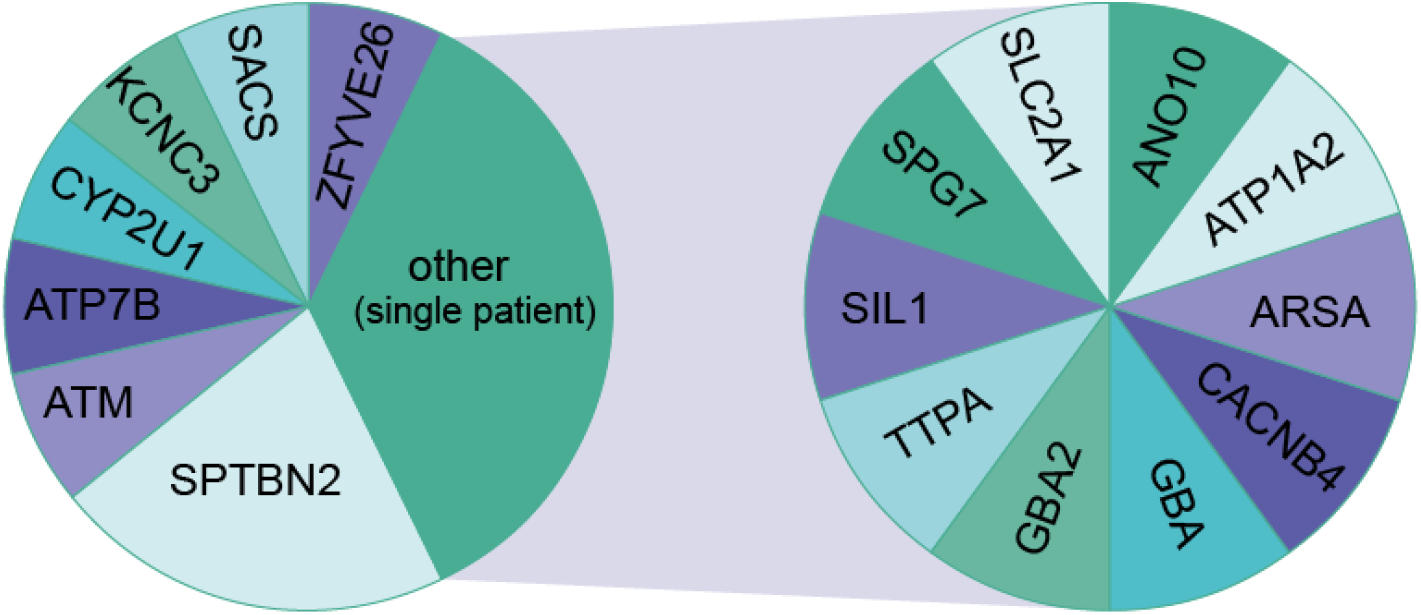
Proportions of genes (single gene and NGS testing) in which DNA variants of uncertain clinical significance were identified

There was a roughly equivalent spread of patients with pathogenic mutations in AR and AD genes (23 vs 21) and a roughly equal number of AR and AD diseases were represented (9 vs 10).

The largest grouping (6/28) of the unclassified variants were identified in *SPTBN2* which is associated with SCA5 (AD) and SCA14 (AR). Single heterozygous variants were found in all individuals. Most were missense but one was an inframe deletion of 2 amino acids. Both forms of SCA can have childhood onset.

Many of the inconclusive results are where a single pathogenic variant has been found in a gene with AR inheritance. The NGS panel testing used in this study will not identify dosage changes in genes and will not detect changes in long range control regions.

## Discussion

MRI is a useful diagnostic tool in children with ataxia as shown by the high rate of abnormal findings (86%). MR spectroscopy (MRS) is routinely used in assessment of adult ataxia patients and during their treatment follow up at the Sheffield Ataxia Centre caring for the adult population (Currie *et al* 2012). Normative data for the paediatric population is now available and will allow future physiological / biochemical assessment of cerebellar function and response to therapy (Balian *et al* - Accepted for poster presentation at ESNR 2022 - September 14-18 Lisbon).

Our data shows a higher diagnostic yield for NGS when compared with adults (45% vs 32%) (Hadjivassiliou *et al* 2018). This is within the expected proportion for children with ataxia of 21-52% identified in previous studies (Subramony *et al* 2022, Krygier and Mazurkiewicz-Beldzinska 2021). It is important to note that the proportion of ataxia resulting from genetic causes is significantly raised in the paediatric population when compared to the adult population. This should be taken into consideration when developing diagnostic and best practice guidelines for paediatric ataxia.

The roughly equivalent rate of patients with AD and AR is a novel finding. Previously it has been a general assumption that diseases with dominant inheritance are responsible for a larger proportion of genetic disease than those with recessive inheritance. This data highlights that both should be considered equally in children with ataxia and this has significant implications for genetic counselling and the risk to siblings.

As seen in adult ataxia data (Hadjivassiliou *et al* 2018), mutations in *FRDA* which is associated with Friedreich’s ataxia and *CACNA1A* which is associated with episodic ataxia type 2 (EA2) are common. Our data places them as the two most common causes of hereditary paediatric ataxia where jointly they represent 43% of positive cases. As expected, the data shows an absence of the late onset diseases seen in the adult data (e.g. SCA6) confirming that there is reduced value in testing ataxic children for SCA’s.

Some of the genes with pathogenic changes or VUS in this population are shared with those seen in previous studies; *SACS, SPG7, KCNC3, TTBK2, CACNA1G, ITPR1, CACNA1A, ATP1A3, SPTBN2, ANO10*. However as with data from multiple previous NGS ataxia cohorts (Subramony *et al* 2022) there are many other genes which are represented by a single case. These rare cases with complex, multisystem phenotypes may not have been diagnosed without a test which covers a comprehensive range of ataxia associated genes. Multi gene panels or whole exome/whole genome sequencing (WGS/WES) are particularly useful for complex paediatric cases with ataxia as diagnoses are often extremely rare and may otherwise require lengthy and costly investigations to determine.

As in the cases with *CACNA1A* pathogenic variants in adult cohorts (Hadjivassiliou *et al* 2018), many of the children who were found to have pathogenic variants in the *CACNA1A* gene were referred for ataxia rather than episodic ataxia. They are therefore likely to be presenting with chronic rather than intermittent ataxia underscoring the importance of inclusion of this gene in ataxia testing and not solely for episodic ataxia testing. This is further supported by the recent expansion of phenotypes reported in the literature to be associated with *CACNA1A*.

The range of disorders associated with *CACNA1A* pathogenic variants was, until recently, believed to be SCA6, episodic ataxia type 2 and familial hemiplegic migraine. Cases have now been reported where pathogenic mutations are present in patients with; congenital ataxia, hemiplegic migraine and developmental delay (Gandini *et al* 2021, Izquierdo-Serra 2020), ischemic stroke with intractable epilepsy and developmental delay (Gudenkauf *et al* 2020), progressive ataxia with hemiplegic migraines or cervical dystonia (Duque *et al* 2021, Fuerte-Hortigon et al. 2020), and at the most severe end of the spectrum cerebral oedema (Gauquelin *et al* 2020). It is possible that increased use of NGS will see congruent phenotype enlargement for other genes with the consequence of overlap into the scope of inherited paediatric ataxia.

The number of *SPTBN2* variants identified both within this study and others is interesting (Subramony *et al* 2022). Pathogenic changes in this gene can be inherited dominantly or recessively and as with *CACNA1A*, the phenotypic spectrum of *SPTBN2* is increasing as a result of NGS. Originally patients identified with a single pathogenic *SPTBN2* variant were found to have late onset progressive ataxia named SCA5. Patients with pathogenic variants on both alleles were found to have congenital or early onset ataxia, named SCAR14 and usually had additional neurological features and intellectual disability. However, it has become apparent that there are many cases of individuals with a single pathogenic missense variant in *SPTBN2* who show similar features to SCAR14 patients (Sancho *et al*. 2021, Nicita *et al*. 2019). Missense changes are commonly identified on NGS screening and are frequently difficult to classify. This is often due to lack of scientific evidence and knowledge surrounding the gene/protein/disorder.

Nearly half (46.7%) of patients where variants were identified using NGS, were reported as inconclusive, as evidence for classification of the variants was limited. Classification of variants found during NGS is complex. Guidelines for interpretation of germline variants are structured to enable evidence from different aspects of the variant to be incorporated into a single numerical score between 1 and 5. 1 represents a benign variant and 5 a variant which is clearly pathogenic. Classification is particularly complex in cases of missense mutations where only a single amino acid has been altered. In such cases the impact of the variant is examined as per the guidelines in UK guidelines for assessment of pathogenicity (Ellard *et al* 2020). This examines the differences between the amino acids which have been exchanged, the position within the protein (i.e. functional domain), conservation of the region, impact on splicing and gene control elements, segregation of symptoms within families or denovo status, functional studies from research settings, matching of diagnostic features present in the patient and frequency within normal control populations. Each category of evidence is weighted depending on the strength of the existing data and these are combined to give an overall classification for the variant.

Standardised DNA variant interpretation is still very much in its infancy. Implementation of current guidelines has been shown to vary between operator with interlaboratory variability at large as 34% (Hoskin *et al* 2017). Additionally, there are many unknowns in terms of impact on regions which are encoded within DNA but which are not traditional protein coding genes such as functional elements and control regions and non-coding RNAs (French *et al* 2020). Even with the best current knowledge, it is frequently not possible to classify a DNA variant.

Many patients in this study were found to have a single pathogenic variant in a recessive gene. Improvements in knowledge of gene control regions and technology allowing screening of long range and intronic control regions and dosage testing may increase the proportion of diagnoses further. However, these additional regions are likely to be small and therefore only account for a low number of cases for the majority of genes. Diagnostic detection rates could be further improved by NGS trio testing. This enables WES or WGS to take place with a greatly reduced burden of data analysis and if used with agnostic gene analysis can identify novel causative genes/ regions. Alternatively, development of functional testing for products of genes with variants of uncertain clinical significance could provide evidence to enable classification. The range of different genes involved makes this complex but it is possible that tests could be developed encompassing several genes with shared cellular pathways.

With the continuing fall in the cost of NGS, the continuing improvements in processing of genomic data due to 100,000 genomes and the standardisation of genetic testing within the NHS, WES/WGS is becoming the standard for rare and complex disease such as paediatric ataxia. Whilst this offers greater overall coverage of the genome, it is important to remember that some regions are poorly covered and this will include genes with known clinical links to ataxia.

## Conclusions

Simple mendelian genetic diseases are responsible for a significant proportion of cases of chronic ataxia in the paediatric cohort.

## Supporting information

Confirmation of ethics approval status from clinical governance

## Data Availability

All data produced in the present study are available in an anonymised format upon reasonable request to the authors

