## Supplementary material for "Genetic Causes of early onset Ataxia: Experience from the National Sheffield Paediatric Ataxia Centre": Confirmation of ethics approval status from clinical governance

Sheffield Children's NHS
NHS Foundation Trust

Western Bank
Sheffield
S10 2TH
www.sheffieldchildrens.nhs.uk

To whom it may concern,

RE: **Publication of - Genetic Causes of Paediatric Ataxia: Experience from the National Specialist Paediatric Ataxia Centre (Dr Santosh Mordekar)**

I am writing to confirm the above project was registered with our clinical governance team at the Sheffield Children's NHS Foundation Trust as a service evaluation project and not a research project. In line with our processes this deemed the project would not require any ethical approval to proceed with the project.

Best wishes

**Keith Pugh**

**Research & Development Manger**

Ruth Brown
Chief Executive

SVS Sheffield Volunteer Standard

Positive About Disabled People

The Children's Hospital Charity

we care

Sarah Jones
Chair
